# Characteristics and needs of Vietnamese technical intern trainees (care workers) in Japan: A qualitative study

**DOI:** 10.1101/2024.05.03.24306822

**Authors:** Koji Hara, Tomokazu Yamamura, Li Ningyi, Pham Thu Huong

## Abstract

**Background:** Foreign caregivers are assuming increasing importance in Japan due to a significant shortage of caregivers. To secure and train stable human resources, it is important to understand the needs and challenges of foreign caregivers.

**Objective:** To identify the needs and challenges of Vietnamese technical interns, the largest group of foreign caregivers in Japan, to facilitate system reforms for foreign caregivers and a more efficient recruitment process.

**Methods:** A semi-structured interview survey was conducted with 27 Vietnamese technical interns (caregivers). Interview items included reasons for choosing technical internship in Japan, desired length of stay, and expectations in Japan, as well as career advancement after returning home. Descriptive statistics and k-means clustering were used to analyze the data.

**Results:** Survey results showed that all respondents had made their own decision to come to Japan and wanted to pursue caregiver certification; 44% had also considered other countries apart from Japan; most wanted to stay in Japan for as long as possible; 37% wanted to live in Japan permanently. The k-means method revealed three clusters: Japanophile cluster or those who preferred Japan for its landscape, culture, and national character; Word-of-mouth cluster or those influenced by personal referrals; and Intellectual cluster or those influenced by Japan’s economic development and care levels.

**Conclusions:** Vietnamese technical interns hoped to study advanced Japanese, become certified care workers, and work in the nursing field after returning to their home country. They also tended to want to stay in Japan for a longer period than that assumed for technical internship programs. Our findings indicate that support for obtaining qualifications, Japanese language skills, and caregiving skills are important to secure stable foreign caregivers. Since each cluster had different backgrounds and reasons, motives, and assumptions for coming to Japan, it is necessary to tailor recruitment, training, and support for each cluster.

## Introduction

Japan’s declining birthrate and aging population are increasing the demand for long-term care; however, there is an acute shortage of care workers to support this demand. According to the Ministry of Health, Labor and Welfare, there were 1.9 million care workers in Japan in 2016, and it is estimated that approximately 2.45 million will be needed by 2045 [1]. The number of care workers has increased by only 15,000 annually, and there are concerns that the workforce will decrease because of the declining birthrate [2]. To address the shortage of care workers, not only do the domestic human resources need to be secured by improving the working conditions and wages of care workers, but the acceptance of foreign care workers also needs to be addressed. In addition new technologies such as information and communication technology and robots need to be utilized [3]. Employment of foreign care workers has been expanding in Japan; according to a survey, 30% of the long-term care facilities in Japan are staffed by foreign care workers [4].

Acceptance of foreign caregivers in Japan may be categorized into the following four types: (1) the Economic Partnership Agreement (the purpose of which is to strengthen bilateral economic partnerships), (2) the status of residence for nursing care (the purpose of which is the acceptance of foreigners in professional and technical fields), (3) the Technical Intern Training Program (the purpose of which is the transfer of skills to their home country), and (4) the Specified Skilled Worker (the purpose of which is the acceptance of foreigners with certain expertise and skills to address labor shortage) [5, 6]. Of the four programs, the Technical Intern Training Program comprises the largest number of foreign caregivers at 7,900, with Vietnam accounting for approximately 40% of the total. However, several challenges have been identified in recruiting foreign caregivers. For example, Yu [7] pointed out that recruitment, training, and traveling costs are incurred as acceptance costs and that a certain level of Japanese language proficiency is required. In addition, they are affected by exchange rates and competition with other countries for human resources [7]. According to the Japan External Trade Organization, the total number of general workers sent abroad from Vietnam is 140,000, with Japan and Taiwan receiving a similar proportion of these workers (approximately 40%), and South Korea (approximately 7%) also growing in popularity as a destination for these workers [8].

Several studies have been conducted on Vietnamese technical trainees coming to Japan [9–12]. However, few studies have focused on their needs, thoughts, and concerns. This information may be useful from the perspective of system reform and human resource recruitment. Therefore, this study conducted an interview survey of Vietnamese technical interns (nurses) prior to their arrival in Japan, regarding their reasons for coming to Japan, their thoughts and concerns about Japan, and their careers after returning home.

## Materials and Methods

### Participant selection and data collection

Technical intern trainees are expected to come to Japan after receiving Japanese language and technical training for several months at the sending agency in Vietnam. Since the regions and characteristics of the participants from each sending agency may be different, this survey was conducted with recruits from three different sending agencies in Hanoi, Vietnam.

The survey was conducted between November 2023 and February 2024 after ethical approval was obtained from the Yokohama City University Ethics Committee (reference number: 2023-18). Participants were sent an explanation of the survey and the survey items in advance. Participants then submitted consent forms to participate.

Semi-structured interviews were conducted with 27 Vietnamese nationals affiliated with Vietnamese sending agencies (nine each from three sending agencies) and scheduled to come to Japan as technical intern trainees (nursing care). Participants were divided into groups of three, for a total of nine groups, and each group was interviewed for approximately 90 minutes by the authors. The interviews were conducted in the conference room of each sending agency, with the assistance of a Vietnamese interpreter. Questions were asked in Japanese and answered in Vietnamese. The survey items are listed in S1 Appendix.

### Data analysis

Descriptive statistics were calculated based on the data obtained from the semi-structured interviews. Clustering using the k-means method with cosine similarity was performed on the 27 participants, who were classified into several clusters. The elbow method was used to determine the number of clusters. The variables used for clustering were selected based on the importance and variance of items. All statistical analyses were performed in R (ver. 4.2.3).

## Results

Participant characteristics are summarized in Table 1. Most participants were female (92.6%). Participants’ average age was 21.7 years, and about half of them (55.6%) were high school graduates, while 22.2% had children. There was no variation in sex or age among the sending agencies, but there was a variation in education and presence of children.

**Table 1.**
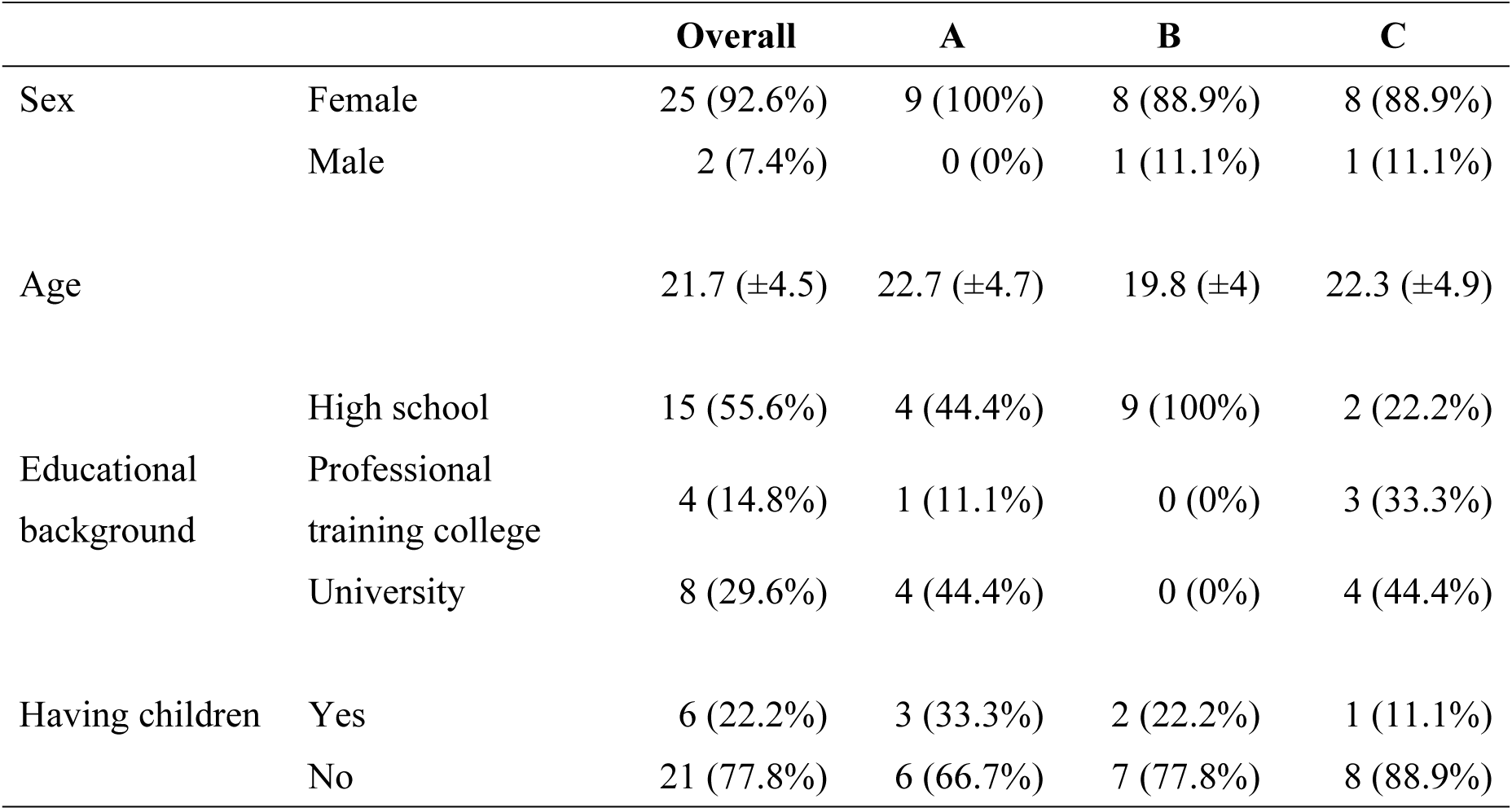
Participants’ Characteristics.

|  |  | Overall | A | B | C |
| --- | --- | --- | --- | --- | --- |
| Sex | Female | 25 (92.6%) | 9 (100%) | 8 (88.9%) | 8 (88.9%) |
|  | Male | 2 (7.4%) | 0 (0%) | 1 (11.1%) | 1 (11.1%) |
| Age | | 21.7 ($\pm 4.5$) | 22.7 ($\pm 4.7$) | 19.8 ($\pm 4$) | 22.3 ($\pm 4.9$) |
| Educational background | High school | 15 (55.6%) | 4 (44.4%) | 9 (100%) | 2 (22.2%) |
|  | Professional training college | 4 (14.8%) | 1 (11.1%) | 0 (0%) | 3 (33.3%) |
|  | University | 8 (29.6%) | 4 (44.4%) | 0 (0%) | 4 (44.4%) |
| Having children | Yes | 6 (22.2%) | 3 (33.3%) | 2 (22.2%) | 1 (11.1%) |
|  | No | 21 (77.8%) | 6 (66.7%) | 7 (77.8%) | 8 (88.9%) |

A, B, and C represent the three sending agencies.

Table 2 shows the interview survey results. More than half the participants (56%) did not consider any other country apart from Japan. Potential destinations other than Japan included Germany (7 respondents), Taiwan (3), Korea (3), and China (1). The reasons for choosing Japan over Germany were the better impression participants had of Japan, the friendly relations between Japan and Vietnam, and the greater availability of information on Japan than on Germany. Taiwan was chosen because of its cultural and lifestyle similarities with Japan. Japan remained the first choice, based on information available on the Internet, and that from acquaintances and relatives in Japan. Korea was less popular because it does not offer nursing care as a career.

**Table 2.** Descriptive Statistics of the Interview Results.

| Questions | Options | n / Average | Percentage / SD |
| --- | --- | --- | --- |
| 1. Whose decision was it to go to Japan? | Self-determined | 27 | 100% |
|  | Other | 0 | 0% |
| 2. Initially, was your family in favor of your going to Japan? | Yes | 20 | 74% |
|  | No | 7 | 26% |
| 3. Did you also consider countries other than Japan? | Yes | 12 | 44% |
|  | Germany | 7 | 26% |
|  | Taiwan | 3 | 11% |
|  | Korea | 3 | 11% |
|  | China | 1 | 4% |
|  | No | 15 | 56% |
| 4. What are your goals in Japan? | JLPT: N2 | 19 | 70% |
|  | JLPT: N1 | 6 | 22% |
|  | Certified Care Worker | 27 | 100% |
| 5. What are your concerns in Japan? | Communication | 17 | 63% |
|  | Living environment/train | 8 | 30% |
|  | Climate (cold) | 5 | 19% |
|  | Disaster | 3 | 11% |
| 6. Do you want to live in Japan permanently? | Yes | 10 | 37% |
|  | No/Undecided | 17 | 63% |
| 7. What is your desired length of stay? | | 6.8 years | $\pm 3.1$ years |
| 8. What do you want to do when you return home? | Care worker | 18 | 67% |
|  | Caregiver Teacher | 5 | 19% |
|  | Other | 4 | 15% |
| 9. How much is your take-home pay in Japan? | | \$1,038.9 | $\pm \$31.5$ |
| 10. What is your desired take-home pay? | | \$1,197.3 | $\pm \$122.9$ |

JLPT, Japanese-Language Proficiency Test. This test has five levels, from N5 to N1, with N1 being the most difficult, and measures the level at which one can understand the Japanese being used in a wide range of situations.
Certified care workers are certified nationally in Japan.
\$1 = 149 yen

Regarding their career goals, 22% of the respondents wanted to pass the most difficult level (N1) of the Japanese-Language Proficiency Test, while all of them wanted to become certified care workers. However, communication difficulties, living environment such as travelling by train, cold weather, and natural disasters, were cited as areas of concern regarding Japan. Only one in three respondents wished to stay in Japan permanently; others wanted to stay for an average of 6.8 years. Many hoped to work in the nursing care industry after returning to their home country.

The reasons for choosing Japan are illustrated in Fig 1. Approximately half of the respondents (51.9%) chose Japan because of their interest in Japanese culture (e.g., anime and kimonos) and scenic beauty (e.g., Mt. Fuji). Approximately 40% of the respondents cited personal recommendation by family and friends and the diligence and rule-abiding nature of Japanese people, respectively, as the reasons for their choice.

**Fig 1.**
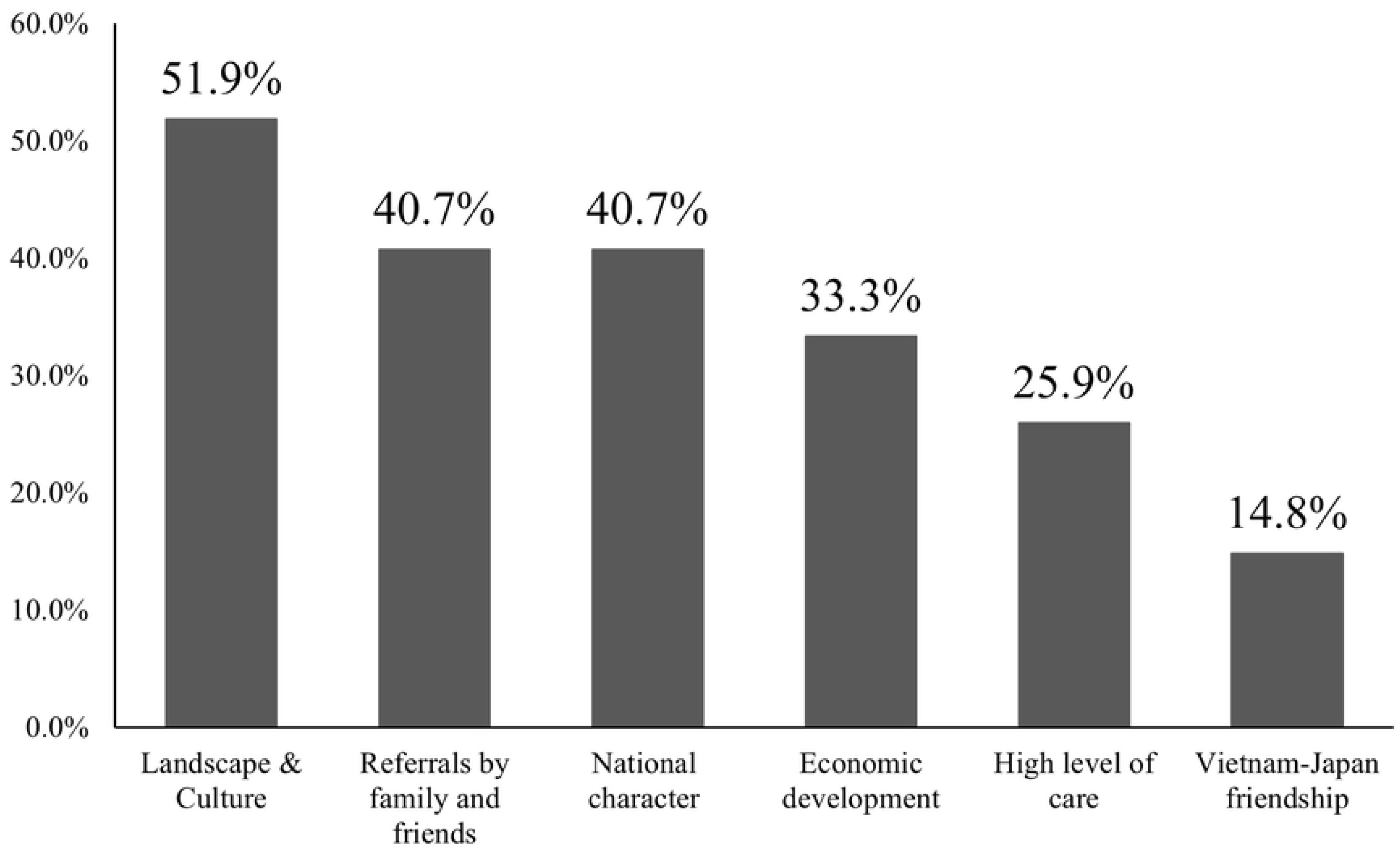
Reasons for Choosing Japan. The number of clusters was determined to be three using the elbow method. The results of clustering using the k-means method are shown in Fig 2 and Table 3. The three clusters were (1) the Japanophile cluster, (2) the word-of-mouth cluster, and (3) the intellectual cluster.

**Fig 2.**
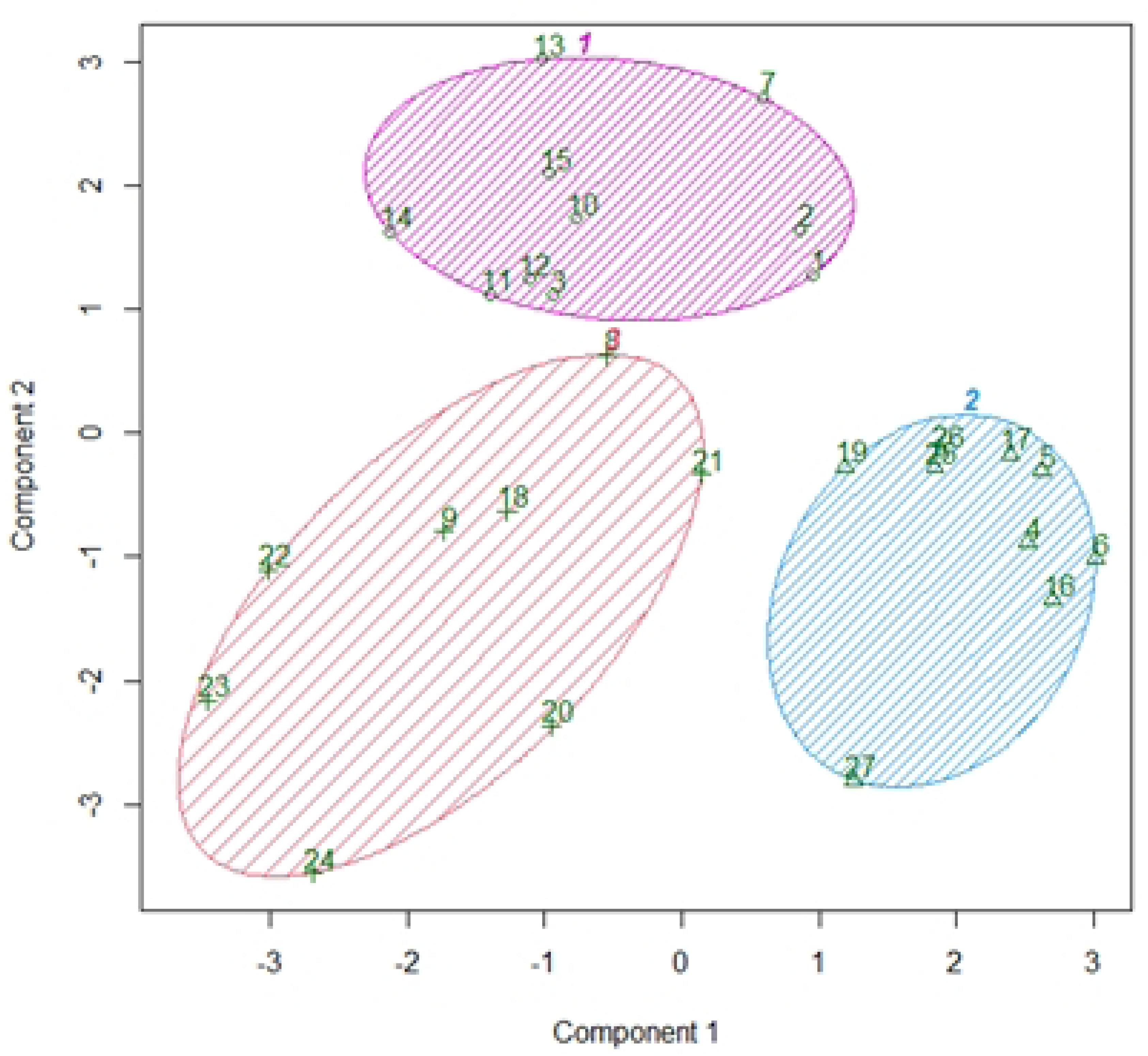
Results of Clustering Using the K-Means Method. Components 1 and 2 explain 38.15% of the point variability.

**Table 3.** Characteristics of the Three Clusters Using the K-Means Method.

| Cluster name | Japanophile | Word-of-mouth | Intellectual |
| --- | --- | --- | --- |
| n | 10 | 9 | 8 |
| Educational background | High school (70.0%)<br>Professional training college (30.0%)<br>University (0%) | High school (55.6%)<br>Professional training college (33.3%)<br>University (11.1%) | High school (37.5%)<br>Professional training college (12.5%)<br>University (50%) |
| Having children | Yes (10.0%)/No (90.0%) | Yes (62.5%)/No (37.5%) | Yes (0%)/No (100%) |
| Main reasons for choosing the technical internship system and sending agency | On-Campus Briefing (80.0%)<br>Referrals by family and friends (30.0%)<br>Internet search (40.0%) | On-Campus Briefing (0.0%)<br>Referrals by family and friends (88.9%)<br>Internet search (33.3%) | On-Campus Briefing (75.0%)<br>Referrals by family and friends (0.0%)<br>Internet search (40.0%) |
| Main reasons for choosing Japan | Landscape & Culture (80.0%)<br>Economic development (70.0%)<br>National character (70.0%) | Referrals by family and friends (66.7%)<br>High level of care (66.7%) | Landscape & Culture (62.5%)<br>Vietnam–Japan friendship (50.0%) |
| Other options for countries | Yes (10.0%, Taiwan)<br>No (90.0%) | Yes (55.6%, Taiwan, Korea, China)<br>No (44.4%) | Yes (75.0%, Germany)<br>No (25.0%) |
| Desire for permanently living in Japan | Yes (10.0%) / No (90.0%) | Yes (33.3%) /No (66.7%) | Yes (75.0%) / No (25.0%) |
| Willingness to take JLPT N1 | Yes (10.0%) /No (90.0%) | Yes (0%) /No (100.0%) | High (62.5%) /No (37.5%) |

The numbers in parentheses indicate the percentage of respondents in the cluster who gave the associated response. JLPT, Japanese Language Proficiency Test.

## Discussion

According to the General Statistics Office of Vietnam, the Vietnamese population aged 65 and above will have grown rapidly, from 7.4 million people in 2019 (7.7% of the population) to 16.8 million people by 2039, and 25.2 million people by 2069 (21.5%) [13]. However, Vietnam has no long-term care insurance and only a few long-term care services. Therefore, family members play an important role in caring for older adults. In recent years, family care has been challenged by declining family sizes due to declining birth rates, a shift from multi-generational cohabitation to nuclear families with an increasing proportion of elderly couples living alone, and rapid urbanization with a large number of rural youths migrating to urban areas, resulting in rapid family dispersal [14]. Consequently, the respondents wanted to gain skills in Japan in anticipation of future care needs or to provide care to their families.

All respondents indicated that they wanted to become certified care workers, which is a Japanese national qualification that effectively allows for permanent residence. However, obtaining this certificate is difficult, with a pass rate of 82.8% for all candidates compared to 43.8% for foreign candidates, under the Economic Partnership Agreement [15]. Although exam-related accommodations have been made for foreign examinees (e.g., paraphrasing of difficult expressions, English notation of diseases, and extended exam times), the learning environment and exam preparation support must be improved. One report also found that gaining a caregiver qualification led to greater understanding of and confidence in their work and contributed to the professionalization of care [16].

In this study, the participants were classified into three clusters. They were divided based on their educational background, reasons for choosing Japan over other candidate countries, and desire for permanent residence. The characteristics of each cluster were then identified. These clusters can be useful for securing and developing human resources in the future. For example, if one wants to target people who wish to live permanently in Japan, targeting the intellectual cluster may be effective for disseminating information via the Internet and emphasizing learning and growth opportunities in Japan. However, when targeting the relatively young age group of high school graduates, it may be prudent to target the Japanophile cluster by visiting high schools to disseminate information emphasizing culture, such as anime and manga, or the diligent and rule-abiding nature of the Japanese people. In addition, when targeting people with high aspirations for caregiving skills, targeting the word-of-mouth cluster may be important by creating a mechanism that fosters referrals for Japanese caregiving. Cluster-based targeting, public relations strategies, and cultivation methods must be given greater consideration while targeting specific care workers.

Japan’s technical internship system is at a major turning point. It has been recommended that the system be revised to protect the human rights of trainees, clarify career advancements, and create a safe, secure, and harmonious society for foreigners [17]. In fact, much research has pointed out problems with the content of work, the treatment of trainees, and the living conditions of technical interns in Japan [11, 18–20]. Many participants in this study were influenced by personal recommendations, particularly regarding the working environment and life in the community. Therefore, promptly establishing a system and providing better working and living environments are among the most important efforts for attracting foreign caregivers.

This study has some limitations that must be considered. First, the participants were limited to 27 individuals from sending agencies in Hanoi; hence, these findings cannot be generalized to all Vietnamese or technical interns. However, by targeting three sending agencies, we avoided significant biases. Second, the participants were interviewed prior to their entry into Japan, and their opinions may change after spending some time in Japan, whereby they may experience a culture shock, conflicting reality or information regarding their assumptions, or a change in their goals and plans; these aspects would need further investigation.

## Conclusions

Our study revealed that Vietnamese technical (nursing) trainees going to Japan wanted to gain advanced Japanese language proficiency and become certified care workers. Most wanted to continue working in the nursing industry after returning to Vietnam. Additionally, more than half the participants chose Japan for care work compared to other countries such as Germany, Taiwan, and Korea, particularly because of Japan’s landscape, culture, and national character, as well as positive referrals by family and friends. More than a quarter of the participants were looking to live in Japan permanently. Given the findings of the clustering of our data, it is important to consider information dissemination, communication, and human resource development tailored to the characteristics of each cluster in order to recruit effectively.

## Data Availability

The datasets generated and/or analyzed during the current study are not publicly available because of individual privacy but are available from the corresponding author upon reasonable request.

## Supporting information

**S1 Appendix. Interview Items.**

## Notes

### Competing Interest Statement

The authors have declared no competing interest.

### Funding Statement

Yes

### Author Declarations

This survey was conducted with the approval of the Yokohama City University Ethics Committee (reference number: 2023-18). During the interview survey, the survey outline was explained to the participants who signed the survey consent form.

